# Healthcare Disparities Among Older Adults: Exploring Social Determinants of Health and Cognition Levels

**DOI:** 10.1101/2024.06.29.24309705

**Authors:** Zahra Rahemi, Juanita-Dawne R. Bacsu, Sophia Z. Shalhout, Maryam S. Sadafipoor, Matthew Lee Smith, Swann Arp Adams

## Abstract

**Background:** The purpose was to investigate the impact of sociodemographic factors on healthcare utilization among adults with different cognition levels (normal and impairment/dementia).

**Methods:** We used cross-sectional data from the Health and Retirement Study (N=17,698) to assess healthcare utilization: hospital stay, nursing home stay, hospice care, and doctor visits.

**Results:** A cohort comparison between normal and dementia/impaired cognition groups revealed significant differences. The dementia/impaired group had lower education levels, higher single/widowed status, and more racial and ethnic minorities. They experienced longer hospital and nursing home stays, varied doctor visit frequencies, and had higher mean age, greater loneliness scores, and lower family social support scores. Differences in hospitalization, nursing home, hospice care, and doctor visits were influenced by factors such as race, age, marital status, education, and rurality.

**Conclusion:** There were disparities in healthcare utilization based on participants’ characteristics and cognition levels, especially in terms of race/ethnicity, education, and rural location.

---

Socio-demographically disadvantaged populations, including racial and ethnic minorities, experience suboptimal care quality and access to healthcare.^1^ Additionally, dementia has emerged as a pressing public health concern. An estimated 6.7 million Americans aged 65 and older live with Alzheimer’s disease and related dementia (ADRD), and this figure is projected to increase to 13.8 million by 2060, driven by growing life expectancy.^2^ In 2023, the estimated financial healthcare cost of dementia in the U.S. was approximately $345 billion, excluding informal caregiving costs.^2^ These observations underscore the critical importance of research efforts to address the mounting challenges associated with healthcare utilization and disparities among diverse older adults and those with cognitive impairment.^3,4^

Socioeconomic factors, such as age, gender, race, ethnicity, education, employment, health literacy, insurance coverage, and the availability of community-based resources, frequently serve as determinants of disparities in access to and utilization of healthcare services.^5–7^ Research indicates that individuals from racial and ethnic minority backgrounds often require more frequent and complex healthcare services, which can potentially result in higher rates of healthcare utilization and increased medical costs.^3,8^ Moreover, Greenwood-Ericksen and Kocher note that rural populations experience an elevated rate of emergency department visits.^9^ The higher risk of preventable emergency department visits, driven partly by a shortage of health professionals, and factors such as delayed care-seeking, cultural norms, and stigma can impede access to proper care, particularly for women.^10^ Limited access to healthcare, lower rates of insurance coverage, and transportation challenges contribute to health disparities in rural areas.^11^ Opportunities for healthy behaviors, including nutritious food consumption, physical exercise, and health promotion, are fewer in these settings. Rural residents often encounter difficulties in reaching healthcare services promptly, and the scarcity of dementia care services accelerates decline. Global shortages in local healthcare providers, telehealth services, and young individuals’ reluctance to stay in rural areas further hinder the addressing of healthcare gaps.^10^ This trend may indicate declining primary care infrastructure, increased care fragmentation, and a deepening healthcare disparity.^9^ Consequently, individuals who are socio-demographically disadvantaged often face poor health outcomes, further exacerbating systemic healthcare disparitie.^8,10^

Healthcare costs for individuals from racial minority backgrounds with ADRD are significantly higher than costs for minority individuals without ADRD as well as costs for White individuals with and without ADRD.^3,12^ It is also reported that older adults with ADRD visit the emergency department more frequently, are hospitalized more frequently, and have longer stays than older adults without ADRD.^13–15^ Recurrent admissions and emergency department visits among individuals with ADRD have been linked to accelerated mortality rates, fall-related injuries, earlier admissions to nursing homes, and cognitive and physical changes.^3,16^

Research is needed to address the growing healthcare challenges and disparities among older adults with diverse socio-demographic identities, especially those with cognitive impairment. While healthcare disparities based on race/ethnicity have been studied, there is a limited exploration of healthcare utilization inequities based on race, ethnicity, and other personal identities concerning ADRD and cognitive disorders.^3^ To gain deeper insight into disparities in ADRD care, we examined healthcare utilization patterns among a national sample of older adults across cognition levels with a focus on key social determinants of health and sociodemographic characteristics, encompassing factors such as education, age, gender, race, ethnicity, marital status, and rural location. We aim to assess the impact of socio-demographic factors on healthcare utilization between individuals with cognitive impairment and those with normal cognition. We used 2014 Health and Retirement Study (HRS) datasets and assessed healthcare utilization using data on the total length of hospital stay, nursing home stay, hospice care, and the number of doctor visits during the last two years.

When assessing disparities in healthcare utilization, especially in relation to sociodemographic and cognitive impairment features, it is important to examine a comprehensive range of service types. Each type of healthcare service provides different insights into the patient’s healthcare needs and usage patterns. By examining all these service types, we aim to understand how sociodemographic and cognitive impairment factors influence overall healthcare utilization. Disparities might not be uniform across different service types. For example, certain sociodemographic groups might have limited access to outpatient care (doctor visits) but may end up having longer hospital admissions due to unmanaged conditions. Similarly, cognitive impairments might lead to increased nursing care stays but not necessarily affect hospice care use in the same way. Further, each service type may be influenced by different barriers (e.g., transportation issues affecting doctor visits). Understanding these can help guide the design of targeted interventions to reduce disparities. Our goal is to identify past trends so that we can compare observations to trends in 2024 and beyond, once the dataset becomes available. This will allow us to assess improvements or lack thereof, and any changes over time in healthcare utilization based on cognition status for different sociodemographic groups. This study provides valuable insights for enhancing the management of older adults’ health conditions, advancing care directives, and addressing current challenges in caring for older adults by focusing on healthcare disparities.

## Materials and Methods

We conducted an observational, cross-sectional investigation using data derived from the Health and Retirement Study (HRS), a nationally representative dataset comprising over 43,000 respondents aged 51 years and older.^17^ The HRS is administered by the Institute for Social Research at the University of Michigan and features biennial data collection across four primary domains: health and well-being, work and retirement, social connections, and economic status. To ensure demographic representation, the survey employs a probability sampling approach, with an oversampling of African American, Hispanic, and Floridian participants. For our study, we analyzed a sample of 17,698 respondents from the HRS 2014 survey (wave 12), focusing particularly on harmonized datasets pertaining to end-of-life topics. Our analysis incorporated data from two HRS sources: the harmonized HRS version B and the 2016 Rand HRS Longitudinal version 2. Since our research involved secondary data analyses, it was deemed exempt from review by [BLINDED] Institutional Review Board.

### Measures

The respondents were classified into two groups using the Langa-Weir approach: dementia/impaired cognition (score of 1-11); and normal cognition (score of 12 or higher).^18^ Given the highly skewed nature of the distributions for each of the healthcare utilization variables (length of hospital stay, nursing home stay, hospice care utilization, and number of doctor visits), these variables were collapsed into four levels: never, low, moderate, and high utilization. A literature search did not reveal any commonly used cut-points. Therefore, we chose cut-points based on natural breaks in the underlying frequency distribution of each variable using a commonly employed data-driven approach where we identified the natural breaks in the data distribution via inspecting histograms or box plots and used gap statistics to determine the optimal number of bins or clusters to help identify significant gaps between clusters. Since the different healthcare services vary in distributions of stay, it is not appropriate to use a single set of cut-points for all variables. For the length of hospitalization and hospice care in the previous two years, the groups were never (0), short (1-7 days), moderate (8-15 days), and long (16+ days). For nursing home care in the previous two years, the points were never (0), short (1-30 days), moderate (31-199 days), and long (120+ days) stays. The number of doctor visits in the previous two years collapsed into never (0), low (1-7 visits), moderate (8-25 visits), and high (26+ visits) number of visits. Other independent variables considered in this study were everyday discrimination, spousal social support, other family social support, and loneliness, which were Likert-type questions in the HRS. However, based on the results of the Wald Chi-Square test from the polytomous logistic regression model, these variables did not achieve statistical significance (p ≥ 0.05) and were consequently excluded from the final regression models. Rurality was determined through a binary question that inquired whether the respondent’s household was in a rural or urban area.

### Statistical Analysis

The analyses were conducted using SAS v9.4 (https://www.sas.com/), and an alpha level of less than or equal to 0.05 was set to determine statistical significance but is exploratory in nature. Due to the different healthcare needs of those individuals with dementia or impaired cognition compared to normal cognition, all analyses were stratified into cognition groups. No features were discovered to be collinear. Given the complex sample design utilized for the HRS, participant sample weights were utilized for all descriptive statistics to allow for nationally representative estimates. Weighted frequencies and means were calculated on variables as appropriate. Weighted sample t-tests and Rao Scott chi-square tests were used to assess for significant differences between cognition groups (dementia/impaired cognition versus normal cognition).

Previous analysis has demonstrated that utilizing the sample weights for multivariable modeling can introduce bias to the estimates;^19^ hence, we did not incorporate these for any model results shown. The Proc logistic function was utilized with the glogit option to conduct polytomous logistic regression models. Each healthcare utilization variable was included as the dependent variable, and demographic variables were incorporated as independent variables. The ‘never’ utilization level was the referent level for each model. All polytomous logistic models were stratified by cognition group (dementia/impaired cognition versus normal cognition) to allow the comparison of different predictors. The p-value for the trend was calculated to determine the significance of the odds ratio decreasing/increasing as utilization level increases across all healthcare types.

## Results

Table 1 describes and compares the demographic characteristics of the normal cognition and dementia/impaired cognition groups from the HRS survey cohort. In contrast to the normal cognition group, the dementia/impaired cognition group participants had lower levels of higher education (61.2% versus 27.2%, p < 0.01) and higher levels of single/widowed status (24.0% vs. 29.6%, p-value < 0.01). Further, more dementia/impaired cognition participants were from racial and ethnic minority communities, including Black and other racial/ethnic groups (p < 0.01) and Hispanic ethnicity (p < 0.01). Compared to the normal cognition group, those in the dementia/impaired cognition group had longer hospital (p < 0.01) and nursing home care stays (p < 0.01) and were significantly more likely to have fewer low and medium number of doctor visits and increased high (26+ visits) as well as no doctor visits at all. In dementia/impaired cognition group, the mean age (p <0.01), loneliness score (p <0.01), and spousal social support score were significantly greater (p <0.01), while the mean score of other family social support was significantly lower (p <0.01). Among the dementia/impaired cognition group, the everyday discrimination score trended higher (p=0.07) than the normal cognition group.

**Table 1.** Descriptive Statistics for the 2014 Heath Retirement Survey Cohort by Cognition Group (HRS, 2014).

|  | Normal Cognition<br>N=13,774<br>Weighted % (n) | Dementia/Impaired Cognition<br>N=3924<br>Weighted % (n) | P-Value |
| --- | --- | --- | --- |
| Education |  |  |  |
| ≤ 12 years | 38.8 (5572) | 72.8 (2733) | < 0.01 |
| 13+ years | 61.2 (7062) | 27.2 (890) |  |
| Gender |  |  |  |
| Male | 45.1 (5304) | 45.4 (1517) | 0.05 |
| Female | 54.9 (7494) | 54.6 (2124) |  |
| Race |  |  |  |
| White | 86.3 (9666) | 66.9 (2084) | < 0.01 |
| Black/Other | 13.7 (2999) | 33.1 (1546) |  |
| Ethnicity |  |  |  |
| Hispanic | 7.1 (1444) | 17.0 (752) | < 0.01 |
| Non-Hispanic | 92.9 (11237) | 83.0 (2882) |  |
| Marital Status |  |  |  |
| Married/Living with a Partner | 76.0 (9743) | 70.4 (2708) | <0.01 |
| Single/Widowed | 24.0 (2952) | 29.6 (931) |  |
| Rurality |  |  |  |
| Rural | 26.4 (3163) | 28.3 (937) | 0.27 |
| Urban | 73.6 (9480) | 71.7 (2678) |  |
| Hospitalization Days Categories |  |  |  |
| None (0 days) | 79.3 (9821) | 70.9 (2514) | <0.01 |
| Low (1-7 days) | 15.7 (2094) | 20.9 (738) |  |
| Medium (7-15 days) | 2.8 (430) | 4.8 (173) |  |
| High (16+ days) | 2.1 (271) | 3.3 (115) |  |
| Hospice Care Days Categories |  |  |  |
| None (0 days) | 58.0 (23) | 49.5 (183) | 0.11 |
| Low (1-7 days) | 21.6 (92) | 23.3 (86) |  |
| Medium (7-15 days) | 9.1 (38) | 9.9 (33) |  |
| High (16+ days) | 11.3 (53) | 17.3 (76) |  |
| Nursing Home Days Categories |  |  |  |
| None (0 days) | 98.0 (12357) | 95.3 (3454) | <0.01 |
| Low (1-30 days) | 1.5 (224) | 3.0 (97) |  |
| Medium (31-119 days) | 0.4 (61) | 1.1 (40) |  |
| High (120+ days) | 0.1 (19) | 0.6 (17) |  |
| Doctors Visits Categories |  |  |  |
| None (0 visits) | 7.6 (1017) | 17.2 (567) | < 0.01 |
| Low (1-6 visits) | 52.1 (6002) | 46.9 (1462) |  |
| Medium (7-25 visits) | 35.6 (4277) | 30.6 (914) |  |
| High (26+ visits) | 4.7 (585) | 5.3 (157) |  |
|  | **Mean (SE)** | **Mean (SE)** |  |
| Age (yrs) | 65.7 (0.3) | 72.3 (0.4) | < 0.01 |
| Everyday Discrimination Score | 1.54 (0.01) | 1.59 (0.02) | 0.07 |
| Spousal Social Support Score | 1.72 (0.01) | 1.86 (0.03) | < 0.01 |
| Other Family Social Support Score | 1.82 (0.01) | 1.76 (0.02) | < 0.01 |
| Loneliness Score | 1.50 (0.01) | 1.63 (0.02) | < 0.01 |

### Hospital Stay

In the normal cognition group, when examining each level of hospitalization, (short, moderate, or long duration stay), compared to those who were never hospitalized, Black individuals were significantly more likely to have moderate hospital stays compared to white participants (p < 0.01). Older age was significantly associated with hospitalization at each level of utilization (p < 0.01). Those individuals who were married were significantly less likely to experience moderate-to-long hospital stays compared to single individuals (p < 0.01). Those who had more than a high school education were less likely to have short or moderate hospital length stays compared to individuals with a high school education or less (p < 0.01, see Table 2- trend test).

**Table 2.**
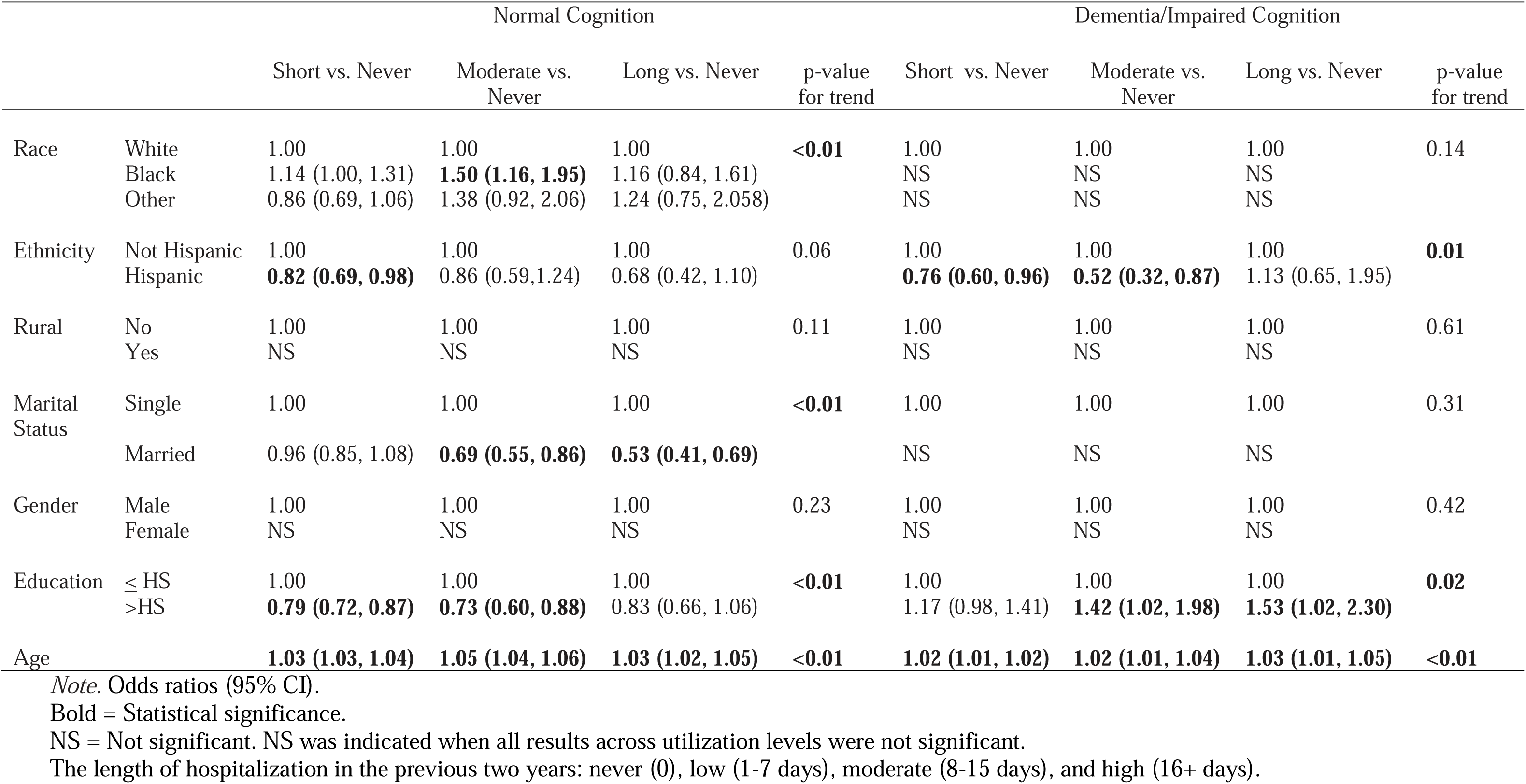
Hospital Days Prediction Model, Health Retirement Survey, 2014.

|  |  | Normal Cognition |  |  |  | Dementia/Impaired Cognition |  |  |  |
| --- | --- | --- | --- | --- | --- | --- | --- | --- | --- |
|  |  | Short vs. Never | Moderate vs. Never | Long vs. Never | p-value for trend | Short vs. Never | Moderate vs. Never | Long vs. Never | p-value for trend |
| Race | White | 1.00 | 1.00 | 1.00 | <**0.01** | 1.00 | 1.00 | 1.00 | 0.14 |
|  | Black | 1.14 (1.00, 1.31) | **1.50 (1.16, 1.95)** | 1.16 (0.84, 1.61) |  | NS | NS | NS |  |
|  | Other | 0.86 (0.69, 1.06) | 1.38 (0.92, 2.06) | 1.24 (0.75, 2.058) |  | NS | NS | NS |  |
| Ethnicity | Not Hispanic | 1.00 | 1.00 | 1.00 | 0.06 | 1.00 | 1.00 | 1.00 | **0.01** |
|  | Hispanic | **0.82 (0.69, 0.98)** | 0.86 (0.59, 1.24) | 0.68 (0.42, 1.10) |  | **0.76 (0.60, 0.96)** | **0.52 (0.32, 0.87)** | 1.13 (0.65, 1.95) |  |
| Rural | No | 1.00 | 1.00 | 1.00 | 0.11 | 1.00 | 1.00 | 1.00 | 0.61 |
|  | Yes | NS | NS | NS |  | NS | NS | NS |  |
| Marital Status | Single | 1.00 | 1.00 | 1.00 | <**0.01** | 1.00 | 1.00 | 1.00 | 0.31 |
|  | Married | 0.96 (0.85, 1.08) | **0.69 (0.55, 0.86)** | **0.53 (0.41, 0.69)** |  | NS | NS | NS |  |
| Gender | Male | 1.00 | 1.00 | 1.00 | 0.23 | 1.00 | 1.00 | 1.00 | 0.42 |
|  | Female | NS | NS | NS |  | NS | NS | NS |  |
| Education | ≤ HS | 1.00 | 1.00 | 1.00 | <**0.01** | 1.00 | 1.00 | 1.00 | **0.02** |
|  | >HS | **0.79 (0.72, 0.87)** | **0.73 (0.60, 0.88)** | 0.83 (0.66, 1.06) |  | 1.17 (0.98, 1.41) | **1.42 (1.02, 1.98)** | **1.53 (1.02, 2.30)** |  |
| Age |  | **1.03 (1.03, 1.04)** | **1.05 (1.04, 1.06)** | **1.03 (1.02, 1.05)** | <**0.01** | **1.02 (1.01, 1.02)** | **1.02 (1.01, 1.04)** | **1.03 (1.01, 1.05)** | <**0.01** |

Note. Odds ratios (95% CI).
Bold = Statistical significance.
NS = Not significant. NS was indicated when all results across utilization levels were not significant.
The length of hospitalization in the previous two years: never (0), low (1-7 days), moderate (8-15 days), and high (16+ days).

Among those with dementia/impaired cognition, when examining each length of hospital stay, those of Hispanic ethnicity were significantly less likely to experience short and moderate lengths of stay compared to the non-Hispanic group (p < 0.01). Individuals with education beyond high school in this group were significantly more likely to experience moderate-to-long hospital stays (p = 0.02). Older age was associated with a significantly increased likelihood of all lengths of hospital stay (p < 0.01).

### Nursing Home Stay

Married individuals with normal cognition, were less likely to utilize nursing homes at any length of time assessed, while older participants were more likely to utilize nursing homes for short, moderate, and long stays (p <0.01). Those in rural areas were significantly less likely to have shorter stays in nursing homes (p <0.01); however, they were significantly more likely to experience longer stays (p <0.01; see Table 3- trend test).

**Table 3.** Nursing Home Days Prediction Model, Health Retirement Survey, 2014.

|  |  | Normal Cognition |  |  |  | Dementia/Impaired Cognition |  |  |  |
| --- | --- | --- | --- | --- | --- | --- | --- | --- | --- |
|  |  | Short vs. Never | Moderate vs. Never | Long vs. Never | p-value for trend | Short vs. Never | Moderate vs. Never | Long vs. Never | p-value for trend |
| Race | White | 1.00 | 1.00 | 1.00 | 0.57 | 1.00 | 1.00 | 1.00 | <0.01 |
|  | Black | NS | NS | NS |  | 0.63 (0.38, 1.04) | 0.58 (0.27, 1.24) | **0.22 (0.10, 0.48)** |  |
|  | Other | NS | NS | NS |  | 1.31 (0.61, 2.79) | 0.79 (0.18, 3.48) | 0.84 (0.29, 2.39) |  |
| Ethnicity | Not Hispanic | 1.00 | 1.00 | 1.00 | 0.06 | 1.00 | 1.00 | 1.00 | <0.01 |
|  | Hispanic | NS | NS | NS |  | **0.39 (0.19, 0.81)** | **0.22 (0.05, 0.96)** | **0.15 (0.05, 0.51)** |  |
| Rural | No | 1.00 | 1.00 | 1.00 | <0.01 | 1.00 | 1.00 | 1.00 | 0.30 |
|  | Yes | **0.60 (0.43, 0.84)** | 1.21 (0.72, 2.05) | **1.97 (1.10, 3.50)** |  | NS | NS | NS |  |
| Marital Status | Single | 1.00 | 1.00 | 1.00 | <0.01 | 1.00 | 1.00 | 1.00 | <0.01 |
|  | Married | **0.67 (0.49, 0.94)** | **0.41 (0.23, 0.72)** | **0.37 (0.20, 0.70)** |  | 0.70 (0.43, 1.13) | **0.41 (0.20, 0.83)** | **0.50 (0.28, 0.86)** |  |
| Gender | Male | 1.00 | 1.00 | 1.00 | 0.31 | 1.00 | 1.00 | 1.00 | <0.01 |
|  | Female | NS | NS | NS |  | 0.92 (0.72, 1.34) | **2.40 (1.21, 4.76)** | **1.69 (1.08, 2.66)** |  |
| Education | ≤ HS | 1.00 | 1.00 | 1.00 | 0.07 | 1.00 | 1.00 | 1.00 | 0.32 |
|  | >HS | NS | NS | NS |  | NS | NS | NS |  |
| Age |  | **1.08 (1.07, 1.10)** | **1.11 (1.08, 1.14)** | **1.10 (1.07, 1.13)** | <0.01 | **1.04 (1.02, 1.06)** | **1.08 (1.04, 1.11)** | **1.09 (1.07, 1.12)** | <0.01 |

Note. Odds ratios (95% CI).
Bold = Statistical significance.
NS = Not significant. NS was indicated when all results across utilization levels were not significant.
Nursing home care in the previous two years: never (0), low (1-30 days), moderate (31-199 days), and high (120+ days).

Among the dementia/impaired cognition and normal cognition groups, marital status (p < 0.01) and age (p < 0.01) had a similar trend effect on nursing home stays: lower utilization for married participants and higher utilization for older individuals. However, in contrast to the normal cognition group, race (p <0.01) and ethnicity (p <0.01) had a significant trend in lower utilization, and females had higher utilization in the dementia/impaired cognition group (p <0.01). Rurality did not have a significant trend. Based upon the point estimate testing, Black participants with dementia/impaired cognition were found significantly less likely to experience long nursing home stays (p <0.01), while those of Hispanic ethnicity were less likely to utilize nursing homes for short, moderate, or long-term care (p <0.01).

### Hospice Care

Among the normal cognition group, the point estimate analysis indicated that those in rural areas were significantly less likely to experience a short length of stay than those in urban areas (p = 0.04) (Table 4). Among those with dementia/impaired cognition, Black individuals were less likely to have short and moderate utilization compared to White individuals (p =0.03). Furthermore, with increasing age (p <0.01), individuals with impaired cognition or dementia were more likely to utilize hospice care (short, moderate, and long durations) (see Table 4- trend test).

**Table 4.** Hospice Care Prediction Model, Health Retirement Survey, 2014.

|  |  | Normal Cognition |  |  |  | Dementia/Impaired Cognition |  |  |  |
| --- | --- | --- | --- | --- | --- | --- | --- | --- | --- |
|  |  | Short vs. Never | Moderate vs. Never | Long vs. Never | p-value for trend | Short vs. Never | Moderate vs. Never | Long vs. Never | p-value for trend |
| Race | White | 1.00 | 1.00 | 1.00 | 0.80 | 1.00 | 1.00 | 1.00 | **0.03** |
|  | Black | NS | NS | NS |  | **0.45 (0.22, 0.93)** | **0.09 (0.01, 0.72)** | 0.86 (0.44, 1.68) |  |
|  | Other | * | * | * |  | NS | NS | NS |  |
| Ethnicity | Not Hispanic | 1.00 | 1.00 | 1.00 | 0.24 | 1.00 | 1.00 | 1.00 | 0.73 |
|  | Hispanic | NS | NS | NS |  | NS | NS | NS |  |
| Rural | No | 1.00 | 1.00 | 1.00 | **0.04** | 1.00 | 1.00 | 1.00 | 0.56 |
|  | Yes | **0.48 (0.27, 0.85)** | 0.83 (0.39, 1.78) | 0.52 (0.25, 1.07) |  | NS | NS | NS |  |
| Marital Status | Single | 1.00 | 1.00 | 1.00 | 0.10 | 1.00 | 1.00 | 1.00 | 0.25 |
|  | Married | NS | NS | NS |  | NS | NS | NS |  |
| Gender | Male | 1.00 | 1.00 | 1.00 | 0.17 | 1.00 | 1.00 | 1.00 | 0.47 |
|  | Female | NS | NS | NS |  | NS | NS | NS |  |
| Education | ≤ HS | 1.00 | 1.00 | 1.00 | 0.40 | 1.00 | 1.00 | 1.00 | 0.63 |
|  | > HS | NS | NS | NS |  | NS | NS | NS |  |
| Age |  | NS | NS | NS | 0.33 | **1.03 (1.01, 1.06)** | **1.08 (1.03, 1.13)** | **1.03 (1.01, 1.06)** | **<0.01** |

Note. Odds ratios (95% CI).
Bold = Statistical significance.
NS = Not significant. NS was indicated when all results across utilization levels were not significant
\* = For the normal models, the black and other groups were combined due to a zero cell count for the other category.
The length of hospice care in the previous two years: never (0), low (1-7 days), moderate (8-15 days), and high (16+ days)

### Doctor Visits

Among the normal cognition group, ethnicity (p <0.01), rurality (p =0.02), marital status (p <0.01), gender (p <0.01), education (p <0.01), and age (p <0.01) had a significant trend across the number of doctor visits (Table 5- trend test). Among the dementia/impaired cognition group, race (p <0.01), ethnicity (p <0.01), rurality (p=0.04), gender (p <0.01), education (p <0.01), and age (p =0.02) had significant trends in the number of doctor visits. For both groups, female gender, higher education, and age were associated with higher doctor visits at all levels. Ethnicity had a similar effect in both groups, where Hispanic participants were shown to be less likely to visit the doctor at all levels. Black individuals with dementia/impaired cognition were less likely to have a moderate-to-high number of doctor visits compared to the white participants. In the dementia/impaired cognition group, those residing in rural areas were only less likely to utilize a high number of doctor visits compared to those living in urban areas. The p-value for the trend for rurality was statistically significant among both normal and dementia/impaired groups (odds ratios decrease as utilization level increases). In both groups, women, older individuals, and those with higher education were more likely to have more doctor visits. In the normal cognition group, those who were married had more doctor visits (Table 5- trend test).

**Table 5.** Doctor’s Visits Prediction Model, Health Retirement Survey, 2014.

|  |  | Normal Cognition |  |  |  | Dementia/Impaired Cognition |  |  |  |
| --- | --- | --- | --- | --- | --- | --- | --- | --- | --- |
|  |  | Low vs. Never | Moderate vs. Never | High vs. Never | p-value for trend | Low vs. Never | Moderate vs. Never | High vs. Never | p-value for trend |
| Race | White | 1.00 | 1.00 | 1.00 | 0.11 | 1.00 | 1.00 | 1.00 | <0.01 |
|  | Black | NS | NS | NS |  | 0.82 (0.64, 1.06) | **0.76 (0.58, 0.99)** | **0.37 (0.23, 0.59)** |  |
|  | Other | NS | NS | NS |  | 0.94 (0.69, 1.29) | 0.90 (0.63, 1.29) | 1.00 (0.55, 1.80) |  |
| Ethnicity | Not Hispanic | 1.00 | 1.00 | 1.00 | <0.01 | 1.00 | 1.00 | 1.00 | <0.01 |
|  | Hispanic | **0.32 (0.27, 0.38)** | **0.23 (0.19, 0.28)** | **0.28 (0.19, 0.39)** |  | **0.34 (0.26, 0.45)** | **0.25 (0.19, 0.35)** | **0.22 (0.13, 0.37)** |  |
| Rural | No | 1.00 | 1.00 | 1.00 | **0.02** | 1.00 | 1.00 | 1.00 | **0.04** |
|  | Yes | 1.14 (0.97, 1.35) | 1.02 (0.86, 1.21) | 0.93 (0.73, 1.20) |  | 0.86 (0.68, 1.08) | 0.81 (0.63, 1.04) | **0.53 (0.34, 0.83)** |  |
| Marital Status | Single | 1.00 | 1.00 | 1.00 | <0.01 | 1.00 | 1.00 | 1.00 | 0.06 |
|  | Married | **1.61 (1.39, 2.03)** | **1.36 (1.17, 1.58)** | 1.06 (0.95, 1.33) |  | NS | NS | NS |  |
| Gender | Male | 1.00 | 1.00 | 1.00 | <0.01 | 1.00 | 1.00 | 1.00 | <0.01 |
|  | Female | **1.56 (1.37, 1.77)** | **1.90 (1.65, 2.17)** | **1.66 (1.35, 2.03)** |  | **1.33 (1.09, 1.61)** | **1.55 (1.25, 1.92)** | **2.05 (1.42, 2.96)** |  |
| Education | ≤ HS | 1.00 | 1.00 | 1.00 | <0.01 | 1.00 | 1.00 | 1.00 | <0.01 |
|  | >HS | **1.81 (1.58, 2.07)** | **1.94 (1.68, 2.23)** | **1.93 (1.57, 2.37)** |  | **1.46 (1.14, 1.87)** | **1.78 (1.37, 2.32)** | **1.58 (1.05, 2.39)** |  |
| Age |  | **1.02 (1.01, 1.03)** | **1.05 (1.04, 1.05)** | **1.04 (1.03, 1.05)** | <0.01 | 1.01 (1.00, 1.02) | **1.02 (1.01, 1.03)** | 1.01 (0.99, 1.02) | **0.02** |

Note. Odds ratios (95% CI).
Bold = Statistical significance.
NS = Not significant. NS was indicated when all results across utilization levels were not significant
The number of doctor visits in the previous two years: never (0), low (1-76 visits), moderate (8-25 visits), and high (26+ visits).

## Discussion

We assessed disparities in healthcare utilization among a nationally representative and diverse group of older adults with and without cognitive impairment across sociodemographic characteristics. Our findings revealed distinct characteristics between the normal cognition and impaired cognition/dementia groups and substantial variations in their length or frequency of healthcare service utilization. Individuals in the impaired cognition/dementia group were typically older, less educated, single, more often from racial or ethnic minority backgrounds, living alone, and showed higher spousal social support but lower support from other family members. Their everyday discrimination score was marginally higher. Additionally, they tended to utilize a greater proportion of hospital and nursing home care and to have either a high number of doctor visits or no doctor visits, with a lower probability of having low to medium levels of visits. These results may be attributed to factors such as limited access to various healthcare services, disease progression stages, increased long-term care/nursing home utilization and hospitalization (as indicated in our findings), or the presence of comorbidities among individuals with cognitive impairment when compared to those with normal cognition.^2,4,5,10,13^ Conversely, older adults with normal cognition often reside in the community and have moderately frequent visits with healthcare providers rather than opting for long-term care. However, further data analyses are necessary to gain a comprehensive understanding of the contributing factors.

Overall, our findings highlighted significant trends in healthcare utilization: Older age consistently predicted higher healthcare utilization across healthcare service and cognition levels; racial and ethnic minorities with cognitive impairment or dementia tended to utilize healthcare services less frequently; in both cognition groups, marriage was associated with reduced hospital and nursing home stays; and individuals with higher education levels and married status in both groups tended to have more frequent doctor visits. An interesting finding indicated that rural populations use short-term healthcare services (such as nursing homes and hospice care) less frequently but have a higher utilization of longer-term stays. Additionally, they were less likely to have high rates of doctor visits. These rural disparities may be attributed to several factors, including limited access to public transportation, finances, healthcare services, and delayed admissions to nursing homes and other healthcare facilities.^10,20–22^ For instance, limited public transportation and finances can hinder rural residents’ access to doctor visits. Conversely, once rural residents enter long-term care facilities like nursing homes, they often have longer stays. A more in-depth examination and research are necessary to elucidate the precise underlying causes of this pattern.

Our findings were consistent with other researchers indicating that healthcare disparities persist in relation to variations in socioeconomic factors, particularly with respect to age, race/ethnicity, education, and rural location.^1,23–25^ Charron-Chenier and Mueller found that despite having greater clinical needs, households who were Black consistently demonstrated lower levels of healthcare use, even when controlling for socioeconomic, demographic, and healthcare insurance coverage differences between groups,^8^ which underscores the existence of health disparities in both health outcomes and access to healthcare.

Our findings confirmed the results of other studies that older adults from racial and ethnic minority backgrounds^3,8^ and rural populations^9,10^ and those with cognitive impairment^13–15^ may use different types and frequencies of healthcare services, potentially leading to increased healthcare utilization rates and costs among these demographic groups. Healthcare costs for racial minorities with ADRD are significantly higher than for minority individuals without ADRD and White individuals, regardless of their ADRD status.^3,12^ Concerning healthcare utilization among racial/ethnic minority individuals with ADRD, certain cultural factors, such as extended family support and the presence of younger caregivers, can have beneficial impacts,^3^ such as decreasing the rate and length of nursing home and hospital stays. However, systemic factors, such as disparities in education and limited access to insurance, may render minority populations more vulnerable, especially in the context of dementia care. Our study aligns with existing research that highlights the presence of healthcare disparities rooted in sociodemographic factors across the U.S.^9,26^ These disparities can have adverse effects on the health of individuals from various communities, especially racial/ethnic minorities and rural populations.

### Strengths and Limitations

A notable advantage of this study lies in its utilization of a substantial national sample comprising older adults across the U.S. Nonetheless, it is important to acknowledge certain limitations that have implications for the interpretation of the findings. First, secondary data analysis is inherently restricted by the data and measurement tools provided within the dataset, over which researchers have no direct influence. Second, the HRS study design results in an overrepresentation of African Americans, Hispanics, and Florida residents within this dataset. To maintain the integrity of our analysis, we purposefully employed survey weights for descriptive statistics, while opting not to incorporate them in the modeling analyses. This approach was selected to ensure the provision of the most precise and unbiased estimates of the associations, aligning with established practices in the academic literature.

### Implications

The implications of our findings extend to research, policy, and practice. By exploring the potential factors contributing to healthcare utilization, we can inform interventional research focused on reducing healthcare disparities. Furthermore, our results hold significance in the development of culturally tailored care management programs and policies that address the distinct requirements of diverse groups and enhance the allocation of healthcare resources in dementia care and advance care planning. To design healthcare for the diverse population of older adults, understanding their values and desires and respecting their sense of dignity is crucial.^27^

## Conclusion

Our findings indicated and confirmed that there were disparities in healthcare utilization based on individuals’ sociodemographic characteristics. Interestingly, we demonstrate utilization is also influenced by cognition levels. This study showed the impact of multiple factors, including age, gender, race, ethnicity, education, marital status, and rural location, on healthcare utilization in individuals with different cognition levels. Our findings on healthcare utilization and disparities in dementia care offer valuable insights that align with the focus on practical advice for managing acute and chronic disorders, advancing care directives, and addressing current issues in the long-term care of older adults. This research can inform the development of culturally tailored care management programs and policies, enhancing the allocation of healthcare resources and supporting the diverse needs and dignity of older adults. Research is needed to guide the design of care planning interventions that support diverse older adults, addressing their healthcare needs and respecting their values and wishes.

## Data Availability

All data produced in the present work are contained in the manuscript.

